# Evaluation of serological lateral flow assays for severe acute respiratory syndrome coronavirus-2

**DOI:** 10.1101/2021.01.02.20248998

**Authors:** Bianca A. Trombetta, Savannah E. Kandigian, Robert R. Kitchen, Korneel Grauwet, Pia Kivisäkk Webb, Glenn A. Miller, Charles G. Jennings, Sejal Jain, Samara Miller, Yikai Kuo, Thadryan Sweeney, Tal Gilboa, Maia Norman, Daimon P. Simmons, Christopher E. Ramirez, Melissa Bedard, Catherine Fink, Jina Ko, Esmarline J. De León Peralta, Gerald Watts, Emma Gomez-Rivas, Vannessa Davis, Rocky Barilla, Jianing Wang, Pierre Cunin, Samuel Bates, Chevaun Morrison-Smith, Benjamin Nicholson, Edmond Wong, Leena El-Mufti, Michael Kann, Anna Bolling, Brooke Fortin, Hayden Ventresca, Wen Zhou, Santiago Pardo, Megan Kwock, Aditi Hazra, Leo Cheng, Q. Rushdy Ahmad, James A. Toombs, Rebecca Larson, Haley Pleskow, Nell Meosky Luo, Christina Samaha, Unnati M. Pandya, Pushpamali De Silva, Sally Zhou, Zakary Ganhadeiro, Sara Yohannes, Rakiesha Gay, Jacqueline Slavik, Shibani S. Mukerji, Petr Jarolim, David R. Walt, Becky C. Carlyle, Lauren L. Ritterhouse, Sara Suliman

## Abstract

**Background:** COVID-19 has resulted in significant morbidity and mortality worldwide. Lateral flow assays can detect anti-Severe Acute Respiratory Syndrome Coronavirus-2 (SARS-CoV-2) antibodies to monitor transmission. However, standardized evaluation of their accuracy and tools to aid in interpreting results are needed.

**Methods:** We evaluated 20 IgG and IgM assays selected from available tests in April 2020. We evaluated the assays’ performance using 56 pre-pandemic negative and 56 SARS-CoV-2-positive plasma samples, collected 10-40 days after symptom onset, confirmed by a molecular test and analyzed by an ultra-sensitive immunoassay. Finally, we developed a user-friendly web app to extrapolate the positive predictive values based on their accuracy and local prevalence.

**Results:** Combined IgG+IgM sensitivities ranged from 33.9% to 94.6%, while combined specificities ranged from 92.6% to 100%. The highest sensitivities were detected in Lumiquick for IgG (98.2%), BioHit for both IgM (96.4%), and combined IgG+IgM sensitivity (94.6%). Furthermore, 11 LFAs and 8 LFAs showed perfect specificity for IgG and IgM, respectively, with 15 LFAs showing perfect combined IgG+IgM specificity. Lumiquick had the lowest estimated limit-of-detection (LOD) (0.1 μg/mL), followed by a similar LOD of 1.5 μg/mL for CareHealth, Cellex, KHB, and Vivachek.

**Conclusion:** We provide a public resource of the accuracy of select lateral flow assays with potential for home testing. The cost-effectiveness, scalable manufacturing process, and suitability for self-testing makes LFAs an attractive option for monitoring disease prevalence and assessing vaccine responsiveness. Our web tool provides an easy-to-use interface to demonstrate the impact of prevalence and test accuracy on the positive predictive values.

## Introduction

Coronavirus disease 2019 (COVID-19), caused by infection with the severe acute respiratory syndrome coronavirus 2 (SARS-CoV-2), was declared a global pandemic on March 11^th^, 2020^1^. A second wave of the pandemic is well underway^1, 2, 3, 4^. However, accurate estimates of transmission rely on accurate and widely available immunosurveillance tools to measure SARS-CoV-2 infection in diverse community settings. Among SARS-CoV-2-infected individuals, 40-45% are estimated to remain asymptomatic^5^, suggesting that prevalence is likely underestimated^6^. Therefore, detecting prior exposure to SARS-CoV-2 as opposed to other viruses, including other coronaviruses is crucial^7^.

There are different types of clinical SARS-CoV-2 tests^8^. Diagnostic testing relies on reverse-transcriptase quantitative polymerase chain reaction (RT-qPCR) and antigen-based immunodiagnostics to detect active infection^9, 10, 11^. Conversely, serological tests can monitor population prevalence and prior exposure by measuring antibodies against SARS-CoV-2^12, 13, 14^. These include enzyme-linked immunosorbent assays (ELISAs), chemiluminescence assays, and lateral flow assays (LFAs)^11, 15, 16^. LFAs are attractive for home testing and population surveillance, since they are affordable, scalable, rely on easily accessible specimens such as fingerstick whole blood and give a result readout within minutes^13^. Since multiple vaccines received emergency use authorization^17^, LFAs could be used to determine whether vaccines elicit a detectable and durable immune response^18, 19, 20, 21, 22^. Furthermore, early seroconversion was shown to predict better clinical outcomes^23, 24^. Hence, easy-to-use LFAs will have important applications in the upcoming phases of the pandemic. Since the onset of the COVID-19 epidemic, multiple studies evaluated the accuracy of serological tests^15, 25, 26, 27, 28, 29^. Many of these tests received Emergency Use Authorization (EUA) through the Food and Drug Administration (FDA)^30^.

Despite the utility of SARS-CoV2 antibody tests, misinterpretation of results is very likely^31^. A negative serological test result does not preclude prior infection since seroconversion occurs 9-11 days after symptoms onset for IgM and 18-20 days for IgG antibodies, respectively^16, 32, 33, 34^. Conversely, positive results do not indicate active infection^31^. Furthermore, the prevalence of SARS-CoV-2 is highly variable ^1, 6^, and known to directly impact the predictive value of a test result. A higher prevalence increases the likelihood that positive test results indicate a real infection (i.e. higher positive predictive value)^35^, but will also decrease the negative predictive value, resulting in more false negative results^35^. Therefore, accessible tools to assist the public with interpreting results based on test accuracy and different prevalence scenarios are critical^31, 36^.

In April 2020, the Mass General Brigham Center (MGB) for COVID Innovation’s direct-to-consumer working group scanned available serological assays and selected 20 lateral flow assays, based on reported assay characteristics and supply chain availability^37^. The LFAs were evaluated by blinded operators using the same samples to standardize the evaluation of their accuracy. Additionally, we developed a user-friendly web-tool to provide context for the end user to interpret their results. Here, we report the evaluation data to serve as a public resource to guide implementation of LFAs, and the tool to aid the interpretation of home testing results.

## Methods

### Sample procurement

We procured 56 SARS-CoV-2 positive, 46 pre-pandemic negative, and 10 HIV+ EDTA plasma samples. SARS-CoV-2 positive EDTA plasma samples were obtained from clinical discards banked within 24-72 hours of collection at the Crimson Core of the MGB Biobank from hospitalized symptomatic patients. All samples had positive SARS-CoV-2 PCR results using an EUA approved test at the Brigham and Women’s Hospital (Panther Fusion SARS-CoV-2 assay, Hologic, Inc., San Diego, CA or Xpert Xpress SARS-CoV-2, Cepheid, Inc., Sunnyvale, CA) or the Clinical Research Sequencing Platform at the Broad Institute of MIT and Harvard (in house Laboratory Developed Test) 10-40 days prior to sample collection. The participants’ charts were reviewed by study staff to identify samples collected ≥10 days after onset of symptoms and to exclude immunosuppressed participants, after which samples were anonymized and stripped of protected health information. Pre-pandemic negative control samples were randomly selected from healthy participants with a Charlson Age-Comorbidity Index^38^ score ≤2, with EDTA plasma banked in the MGB Biobank between Jan 1-Dec 1, 2019 from inpatients. HIV-positive control samples were obtained from EDTA plasma samples banked prior to January 2020 in a study on neuropathic pain in HIV. All HIV-positive participants were on antiretroviral therapy. For 8 out of the 10 HIV-positive samples, viral load quantification was available and showed 256 copies/ml or less, and 5 showed either undetectable loads or under 20 copies/ml. The study was approved under the Massachusetts General Brigham (MGB) Institutional Review Board (protocol no. 2020P001204).

### Lateral Flow Assays (LFAs)

Twenty commercial IgG/IgM lateral flow assays (LFAs) from 18 manufacturers were evaluated **(Supplementary Table 1)**. LFAs were analyzed by blinded operators according to manufacturer instructions for use (IFU), with the exception of using micropipettes instead of manufacturer-provided droppers to minimize technical variability. Samples were thawed on ice, randomized, and brought to room temperature. Kit components were also brought to room temperature. The IFU-specified volumes of sample and buffer were added to the cassette. Specified sample volumes varied for different LFAs but were typically in the 5-20μL range. The cassettes were run at room temperature on a flat surface and results read immediately after the time interval defined in the IFU (typically ranging from 10-15 minutes). Each cassette was independently scored by two blinded raters as either “positive,” “negative,” or “invalid”. Ratings were designated according to the interpretation guidelines outlined in each individual IFU. Each cassette was photographed under four standardized illumination conditions and viewpoints for future analysis.

### Reproducibility testing

For inter-operator reproducibility analysis, separate pools of EDTA plasma were obtained from >30 pre-pandemic healthy individuals (negative pool) and >30 convalescent participants collected after symptom resolution at the Massachusetts General Hospital (MGH) respiratory illness clinic (positive pool). Convalescent samples for the positive pool were confirmed to be positive using the COBAS SARS-CoV-2 PCR test (Roche Diagnostics, Indianapolis, IN) at MGH. A total of 20 replicates per pool were run by two independent pairs of blinded operators, alternating between positive and negative pools (10 replicates per pair). Reproducibility was calculated according to agreement between operator ratings as well as concordance of readout with sample pool COVID status.

### Sensitivity and specificity testing

Our cohort of 112 EDTA plasma samples was used across all 20 LFAs to evaluate performance: sensitivity, specificity, positive predictive value (PPV) and negative predictive value (NPV). Samples were sub-aliquoted throughout the analysis to minimize freeze-thaw cycles. Binary presence/absence were used, and discordant calls were resolved by a third operator inspecting photographs taken of the relevant LFA.

### LFA usability

In addition to initial screening ^37^, each LFA kit was assessed for consumer usability based on complexity of kit materials, sample requirements, and IFU clarity. Supplied kit components were documented for completeness and examined for ease-of-use. IFU protocols were rated on a scale from 0-14 according to a predefined rubric **(Supplementary Table 2)** by three independent raters. Usability evaluations are shown in **Supplementary Table 3**. Sample input requirements for each LFA are in **Supplementary Table 1**.

### Ultrasensitive Simoa Serology Assays

Plasma samples were diluted 4000-fold, and the total IgG and IgM levels against the SARS-CoV2 spike protein were measured using a custom Single Molecule Array (Simoa) assay as described^39^ on an automated HD-X Analyzer (Quanterix, Billerica, MA, USA), to provide a quantitative reference for anti-spike antibody titers in the plasma samples. Normalized mean Average Enzymes per Bead (AEB) levels were calculated using a standard set of calibrators produced by serially diluting a large volume of plasma from seroconverted individuals. Antibody concentrations were estimated using a calibration curve of recombinant anti-SARS-CoV-2 antibodies^40^.

### Analysis and Webapp

Detailed methods are in the supplementary.

## Results

### Study Population

We obtained plasma samples from 56 pre-pandemic patients, including 10 HIV+, and 56 symptomatic patients in March and April 2020. HIV-negative samples were matched for sex and age between the COVID- and COVID+ groups. The overall study population included 25.9% Blacks, 4.5% Latinx, 9.8% Asian and 43.8% Non-Hispanic Whites **(Table 1)**. COVID+ samples were from individuals between 10-40 days post symptom onset, with 60.7% samples taken between 2-4 weeks. Of all the COVID+ participants, 7 (12.5%) were symptomatic outpatients and 49 (87.5%) were hospitalized. Among those hospitalized, 25 (51%) required intensive care unit (ICU) treatment and 4 (8.2%) were deceased at the time of chart review **(Table 2)**. Additional mortalities were possible after chart review since some participants were in critical condition in the ICU.

**Table 1:** Demographic information of all individuals whose plasma samples were used for this study. Individuals are broken out by COVID+/- and HIV+/- status. Black includes one mixed African American in the COVID-HIV+ group.

|  | COVID- HIV- | COVID- HIV+ | COVID+ HIV- | Overall |
| --- | --- | --- | --- | --- |
|  | n = 46 | n = 10 | n = 56 | n=112 |
| **Sex** |  |  |  |  |
| Female | 28 (60.9%) | 1 (10%) | 31 (55.4%) | 60 |
| Male | 18 (39.1%) | 9 (90%) | 25 (44.6%) | 52 |
| **Age** |  |  |  |  |
| Mean (SD) | 46.7 (13.4) | 58.6 (5.3) | 58.7 (20.4) | 53.8 (17.8) |
| Median<br>[Min, Max] | 48.0<br>[19.0, 68.0] | 58.0<br>[48.0,65.0] | 57.5<br>[24.0, 98.0] | 54.5 [19.0,98.0] |
| **Race** |  |  |  |  |
| Asian | 8 (17.4%) | 0 (0%) | 3 (5.4%) | 11 (9.8%) |
| Black | 10 (21.7%) | 4 (40%) | 16 (28.6%) | 30 (26.8%) |
| LatinX | 0 (0%) | 0 (0%) | 5 (8.9%) | 5 (4.5%) |
| Unknown/Other | 8 (17.4%) | 1 (10%) | 8 (14.3%) | 17 (15.2%) |
| White | 20 (43.5%) | 5 (50%) | 24 (42.9%) | 49 (43.8%) |

**Table 2:** Clinical information for the COVID-positive individuals whose plasma was used in this study. Individuals are broken out by the number of days between symptom onset and blood collection.

| Interval between symptom onset and blood collection | 7-14 days | 15-21 days | 22-28 days | 29+ days | Overall |
| --- | --- | --- | --- | --- | --- |
|  | n = 9 | n = 18 | n = 16 | n = 13 | n=56 |
| **Sex** |  |  |  |  |  |
| Female | 6 (66.7%) | 10 (55.6%) | 11 (68.8%) | 4 (30.8%) | 31 (55.4%) |
| Male | 3 (33.3%) | 8 (44.4%) | 5 (31.2%) | 9 (69.2%) | 25 (44.6%) |
| **Age** |  |  |  |  |  |
| Mean (Standard Deviation) | 72.0 (24.0) | 53.0 (15.8) | 52.9 (19.9) | 64.7 (20.2) | 58.7 (20.4) |
| Median [Min, Max] | 85<br>[30, 98] | 49<br>[30, 84] | 54.5<br>[24, 88] | 67<br>[29, 94] | 57.5<br>[24, 98] |
| **Race** |  |  |  |  |  |
| Asian | 1 (11.1%) | 1 (5.6%) | 1 (6.2%) | 0 (0%) | 3 (5.4%) |
| Black | 5 (55.6%) | 5 (27.8%) | 2 (12.5%) | 4 (30.8%) | 16 (28.6%) |
| LatinX | 1 (11.1%) | 3 (16.7%) | 1 (6.2%) | 0 (0%) | 5 (8.9%) |
| Unknown/Other | 1 (11.1%) | 1 (5.6%) | 2 (12.5%) | 4 (30.8%) | 8 (14.3%) |
| White | 1 (11.1%) | 8 (44.4%) | 10 (62.5%) | 5 (38.5%) | 24 (42.9%) |
| **Primary COVID PCR test** |  |  |  |  |  |
| Cepheid Xpert Xpress SARS-CoV-2 | 1 (11.1%) | 0 (0%) | 1 (6.2%) | 0 (0%) | 2 (3.6%) |
| Panther Fusion SARS-CoV-2 | 8 (88.9%) | 17 (94.4%) | 12 (75.0%) | 12 (92.3%) | 49 (87.5%) |
| In house LDT at the Broad Institute | 0 (0%) | 1 (5.6%) | 3 (18.8 %) | 1 (7.7%) | 5 (8.9%) |
| **Severity** |  |  |  |  |  |
| Deceased | 0 (0%) | 0 (0%) | 4 (25%) | 0 (0%) | 4 (7.1%) |
| Hospitalized, not ICU | 7 (77.8%) | 13 (72.2%) | 2 (12.5%) | 2 (15.4%) | 24 (42.9%) |
| ICU | 0 (0%) | 0 (0%) | 2 (12.5%) | 2 (15.4%) | 4 (7.1%) |
| ICU (intubated) | 0 (0%) | 1 (5.6%) | 1 (6.2%) | 1 (7.7%) | 3 (5.4%) |
| ICU (intubated) - Recovered | 0 (0%) | 3 (16.7%) | 1 (6.2%) | 7 (53.8%) | 11 (19.6%) |
| ICU (no intubation) - Recovered | 1 (11.1%) | 0 (0%) | 2 (12.5%) | 0 (0%) | 3 (5.4%) |
| Outpatient | 1 (11.1%) | 1 (5.6%) | 4 (25%) | 1 (7.7%) | 7 (12.5%) |

### LFA Performance: Reproducibility

Four independent operators working in teams of two on separate days applied each pool 10 times to each LFA, with processing as dictated by the instructions for use (IFU). API version1 LFAs were not assessed for reproducibility due to limited cassette availability and high sample volumes required. Of the remaining 19 LFAs, three (BTNX, Camtech, and Carehealth) had 100% consistent correct outcomes across both isotypes, with an additional three (BioHit, Zhuhai Livzon, and Phamatech) having no incorrect or inconsistent outcomes with one or two invalid tests **(Figure 1)**. IgG was the more reproducible isotype. The majority of incorrect consistent calls came from operators calling a COVID+ sample IgM negative **(Figure 1)**.

**Figure 1:**
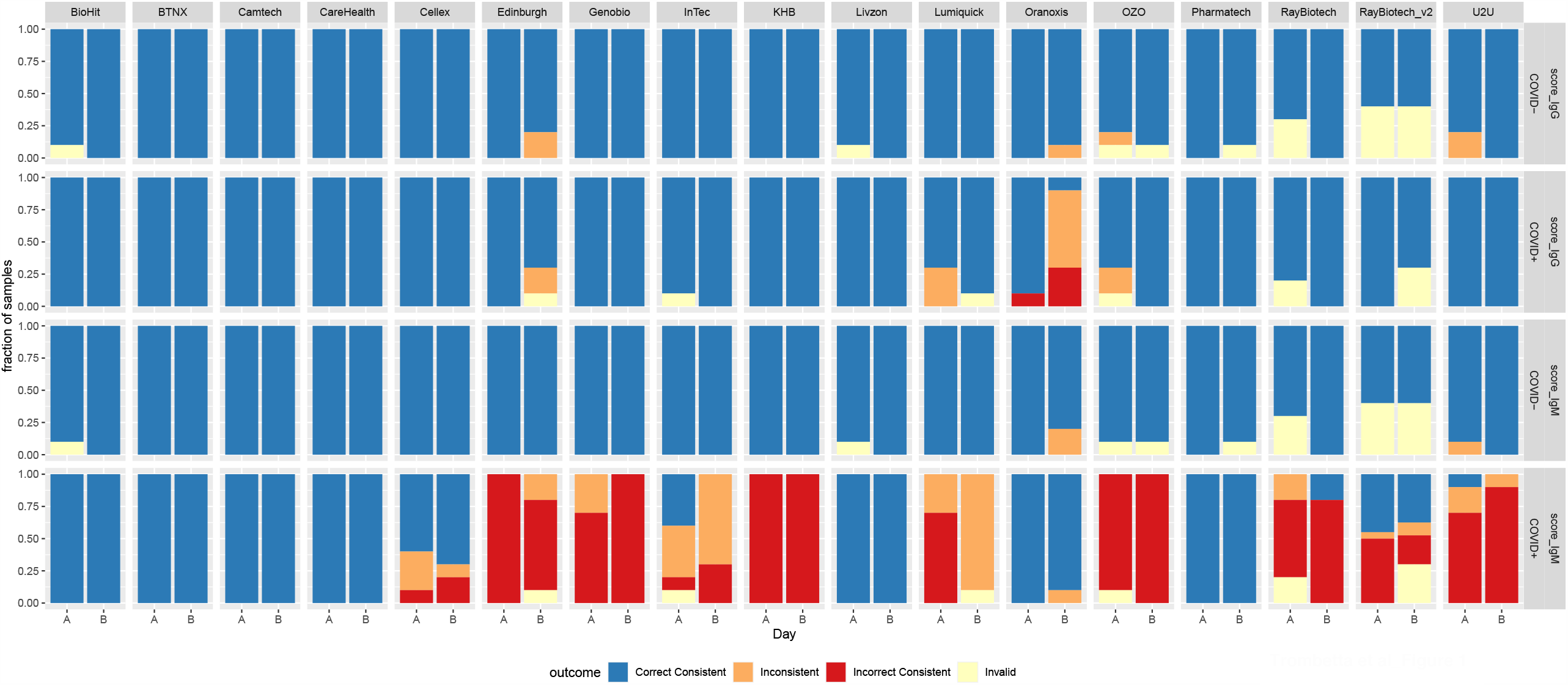
LFA reproducibility. Colors represent LFA outcome performed across two days (A & B) by two independent pairs of operators on a COVID positive (COVID+) pooled sample and a COVID negative (COVID-) pooled sample. On each day, ten technical replicates of each pool were performed. Blue represents replicates where both operators that day agreed on the outcome, and the outcome was correctly called positive or negative. Red represents occasions where both operators agreed, but the outcome was incorrectly called. Orange represents replicates where the operators did not agree on the LFA outcome. Light yellow represents an invalid test where control bands did not meet criteria for a valid test. IgG was the most reproducible isotype for all LFAs except Oranoxis, and consistent false negatives were common occurrences for IgM for the majority of LFAs.

### LFA Performance: Practicalities of Use

We assessed the LFAs according to this rubric **(Supplementary Table 2)** by three independent raters and assigned a composite score on a scale of 0-14 **(Supplementary Table 3)**. Five LFAs (BioHit, InTec, Lumiquick, Phamatech, and U2U) received full marks for IFU clarity. LFAs frequently lost points for imprecise instructions regarding correct usage of disposable droppers as well as optimal time between adding sample and reading the results. Important kit usability criteria, such as whether the included pipette droppers show clear volume markings, were also recorded **(Supplementary Table 3)**. While these kits may not yet be intended for the general public, it will be important to clarify the instructions moving forward and include clearly marked droppers to minimize potential for sample volume errors.

### LFA Performance: Sensitivity and Specificity

To focus on LFA specificity, given the likely use of these tests in low-prevalence settings, disagreement between operators was interpreted as a negative call. Across all but three of the LFAs (Biohit, BTNX, Vivacheck), sensitivity was higher for IgG than IgM. Sensitivity for IgG ranged from 98.2% (Lumiquick) down to 72.7% (Oranoxis), and for IgM from 96.4% (BioHit) to 23.2% (Oxo and U2U) **(Table 3)**. LFA specificity was much higher for both isotypes, with 11 LFAs having a specificity of 100% for IgG (API, API v2, BTNX, Camtech, Genobio, Oranoxis, Phamatech, Ray Biotech, Ray Biotech_v2, U2U and Zhuhai Livzon), and 8 LFAs having a specificity of 100% for IgM (API, CareHealth, Cellex, Lumiquick, Oranoxis, Ray Biotech v2, U2U, Zhuhai Livzon) **(Table 3)**. Under the assumption that the likelihood of two randomly occurring false positives for any one individual is low, an IgG/IgM composite score (averaged operator scores, see Methods) was produced to maximize test specificity. Using this composite score, all LFAs except BioHit, Cellex, Edinburgh, InTec and Vivachek achieved a specificity of 100%. This result underscores the potential of considering the outcome in both isotypes to minimize false positives, although it is more likely that a single isotype will be used in clinical testing.

**Table 3:**
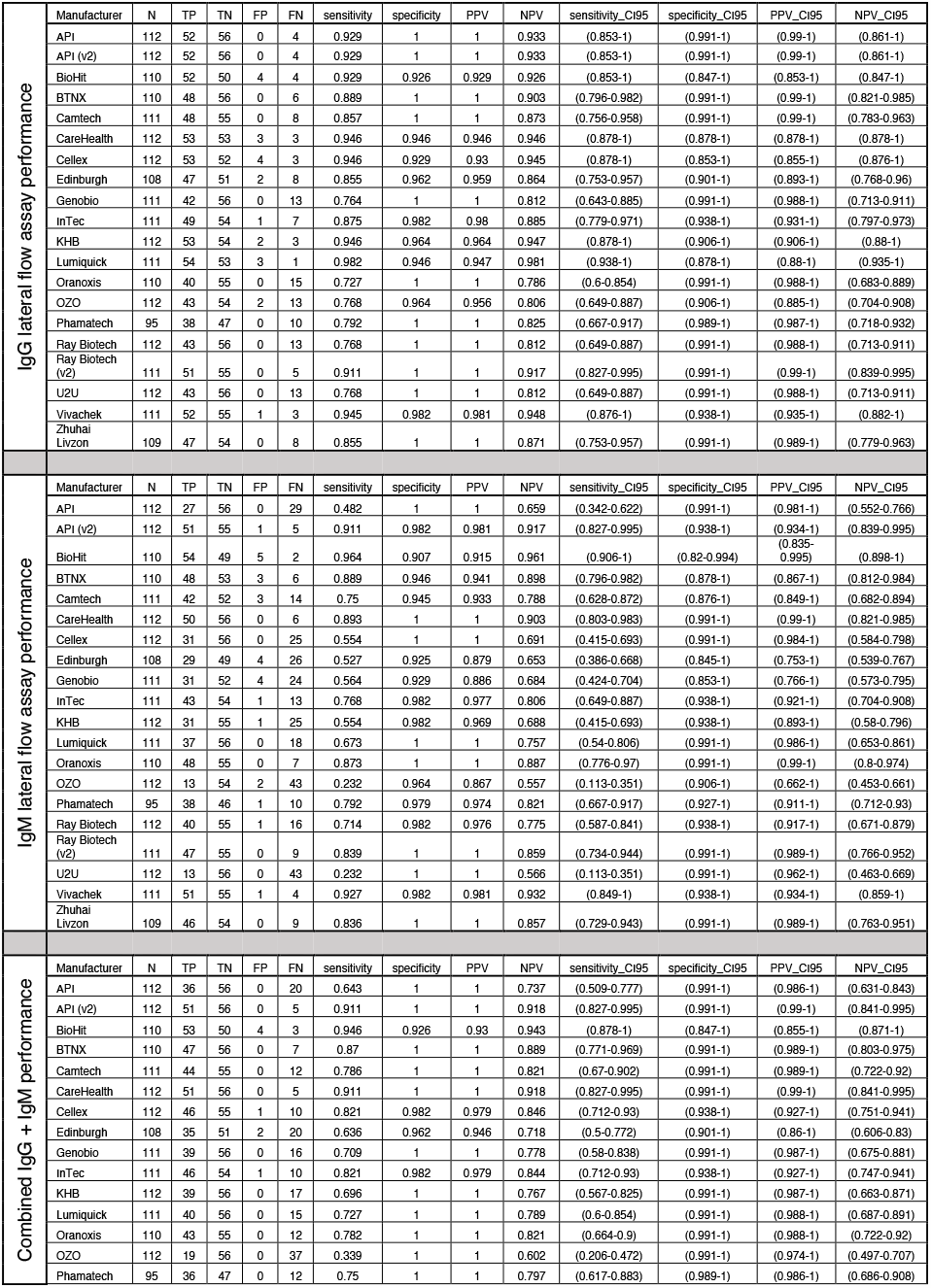
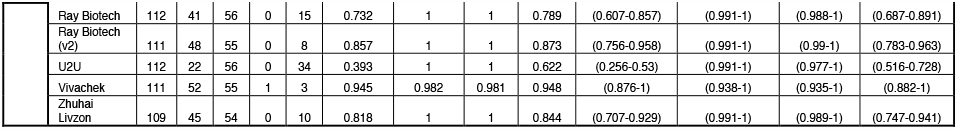
LFA performance in terms of sensitivity and specificity. N: Total number of valid assays; TP: True Positive; TN: True Negative; FP: False Positive; FN: False Negative; PPV: Positive Predictive Value; NPV: Negative Predictive Value; and CI95: 95% confidence interval.

We created heatmaps to visualize individual sample outcomes across all LFAs to assess whether we systematically detected the same miscalls across multiple LFAs **(Figure 2)**. False negatives (blue squares in the COVID+ panel) amongst COVID+ patients were somewhat reproducible, with three COVID+ samples called negative in both isotypes by all or all but one LFA. These miscalls were not clearly explained by known demographics (age, sex) or clinical variables (disease severity, weeks post symptom onset) **(Figure 2**, bottom panel). To investigate whether these miscalls were related to low titers of anti-SARS-CoV-2 antibodies from participants with a suppressed immune response, all 112 samples were analyzed for anti-spike IgG and IgM antibodies using a custom quantitative Simoa assay^39^. The three samples that were called negative across almost all LFAs, which were collected 2-3 weeks after symptoms onset, had the lowest levels of anti-spike antibodies in COVID+ samples for both IgG and IgM, suggesting these participants had slower or suppressed immune responses to SARS-CoV-2 infection.

**Figure 2:**
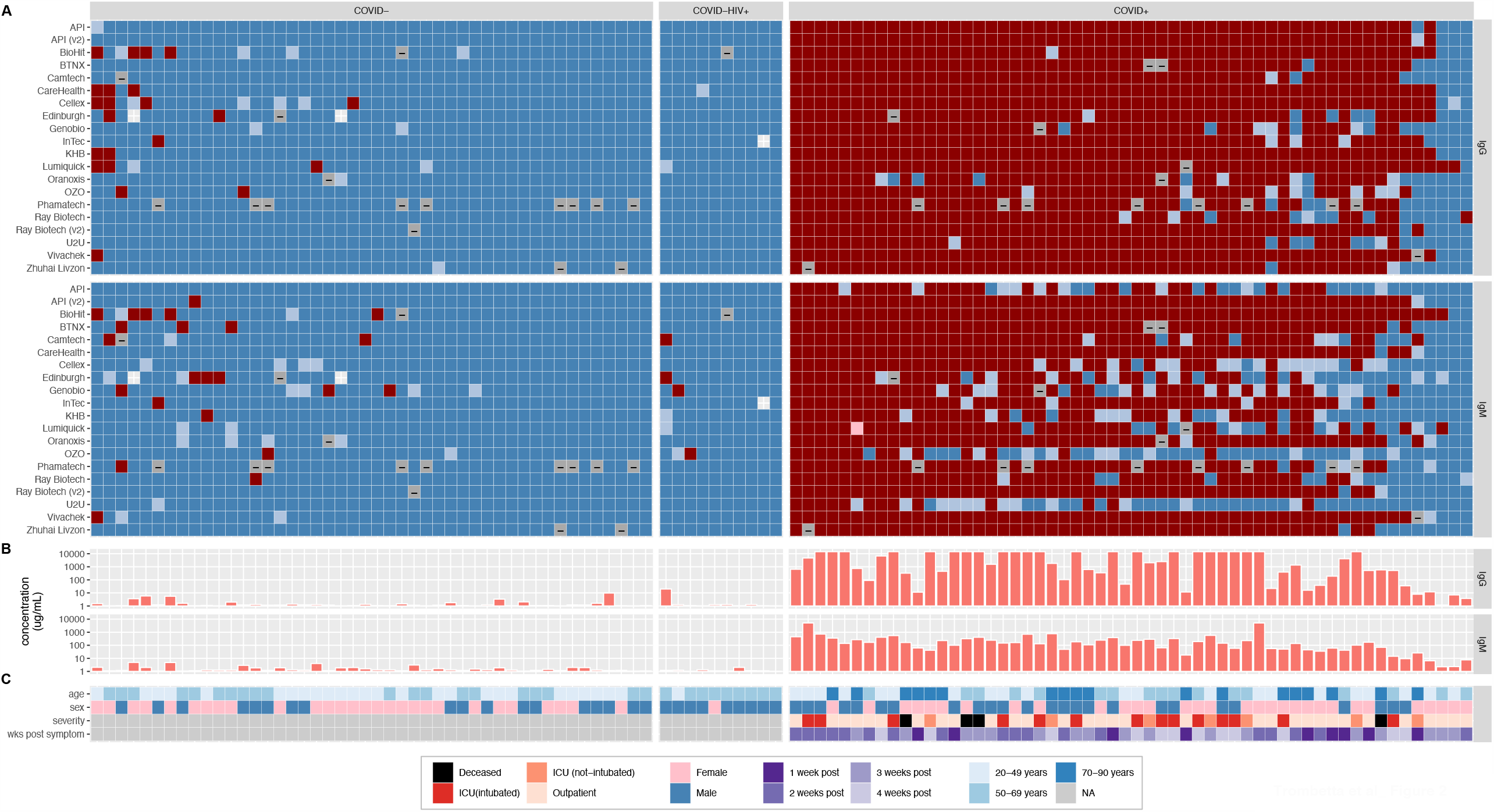
Per-individual LFA performance. Colors represent scores computed for IgG (top panel) and IgM (lower panel), where dark red=+1 (operators all agree a band is present) and dark blue=-1 (operators all agree there is no band) with intermediate colors (pink and light blue) representing varying degrees of operator disagreement. Grey represents invalid runs. Samples are ordered within the COVID-HIV-, COVID-HIV+, and COVID+ (all HIV-) groups in order of decreasing average score across all LFAs for both antibodies. Clinical variables include age (<50yrs: light blue, 50-69yrs: intermediate blue, ≥70yrs: dark blue), sex (blue: male, pink: female), disease severity (hospitalized: light green, ICU: orange, ICU+respirator: red, deceased: black), and weeks post symptom-onset (dark green: 1-2 weeks, lightest green: ≥5 weeks).

Unlike false negatives, most false positives **(Figure 2**, red squares in COVID-panel) amongst COVID-individuals appear largely uncorrelated between LFAs. However, two samples showed IgG false positives across multiple LFAs, which may suggest long lasting antibodies from exposure to coronaviruses other than SARS-CoV-2. Nonetheless, this observation is not reflected in the antibody levels from the Simoa analysis, which showed barely detectable anti-spike antibodies in these two samples.

### Defining the limit of detection for qualitative LFAs

The custom Simoa anti-spike IgG and IgM antibody assays use a standard curve to determine standardized antibody concentration in each sample^39^. The results obtained from this assay can therefore be used to estimate a limit of detection (LOD) for each of the qualitative LFAs. The cumulative number of false negative LFA calls in COVID+ samples **(Figure 3, y-axis)** were computed as a function of decreasing antibody concentrations **(Figure 3, x-axis)** separately for IgG and IgM. We define the LOD for each LFA/antibody as the concentration at which ≥95% of the COVID+ samples are called unambiguously positive **(Supplementary table 4)**. Using this definition, for IgG, all LFAs (except Genobio, Oranoxis, OZO, Ray Biotech, and U2U) have an LOD within the linear range of the SIMOA assay (1 - 10,000 μg/mL). Lumiquick has the lowest LOD at 0.1 μg/mL, which was extrapolated by dilution to be within the linear range of the Simoa standards, and CareHealth, Cellex, KHB and Vivachek all have an LOD of 1.5 μg/mL **(Supplementary table 4)**. Considering the generally lower sensitivity observed with the IgM assays **(Figure 2)**, IgM assays consistently have higher LODs, with 9 exceeding 1,000 μg/mL. BioHit has the lowest IgM LOD at 0.6 μg/mL, and API version2, BTNX, CareHealth and Vivachek all have LODs under 10 μg/mL.

**Figure 3:**
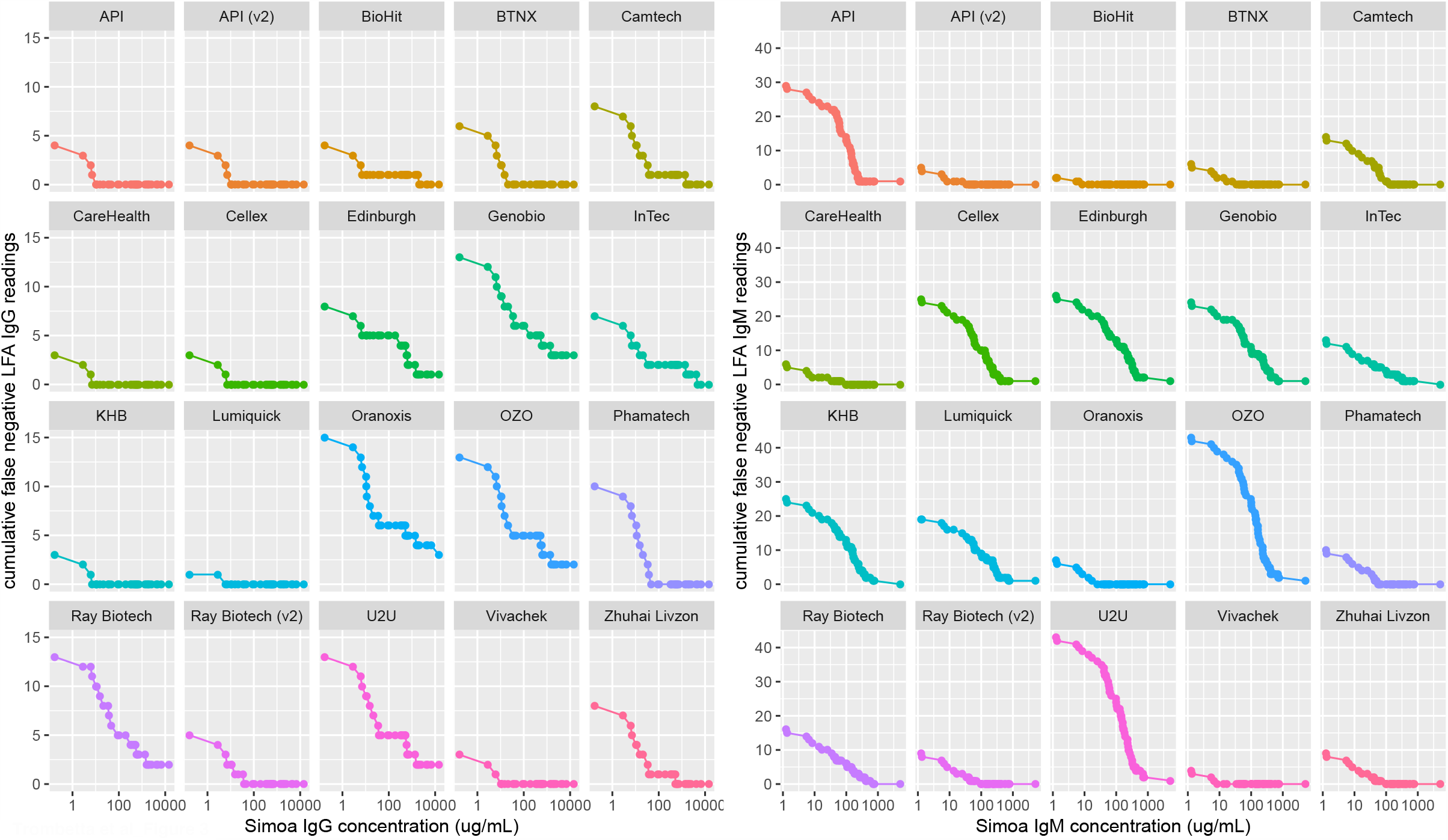
Determining the limit of detection of qualitative LFAs. Samples were ranked from highest concentration of anti-spike antibody (determined by Simoa) on the right, to lowest concentration (x-axis). As the sample concentration decreases to the left, a cumulative count of false negatives is shown on the y-axis. IgG is shown on the left, IgM on the right. Note the difference in magnitude of the y-axes between IgG and IgM; these LFAs are generally more sensitive to IgG than IgM.

### Interpreting test positivity with low prevalence in the general population

Low prevalence places a high burden on specificity^35^. Given the high proportion of true negative individuals in the population being studied, prevalence increases the ratio of false positive to true positive test outcomes^41^. Positive predictive values (PPV) correspond to the likelihood that a positive test result reflect true positivity as measured by a gold standard PCR result. We computed PPVs as a function of the cumulative fraction of the population infected with (and assumed to have produced antibodies to) SARS-CoV-2 **(Figure 4)**. Here we see that even with the conservative interpretation of LFA outcome (requiring the majority of operators to see a band to call a sample positive for either IgG or IgM), when the population seroprevalence is ∼5% the PPV for these assays spans a large range (from ∼30% to 100%). As the population prevalence increases, the burden on specificity is decreased, and at 50% prevalence the PPV of all LFAs is above 87.5%. Posterior PPV can be improved for most LFAs by requiring both IgG and IgM to be read as positive in order to count an individual as positive **(Figure 4, right panels)**.

**Figure 4:**
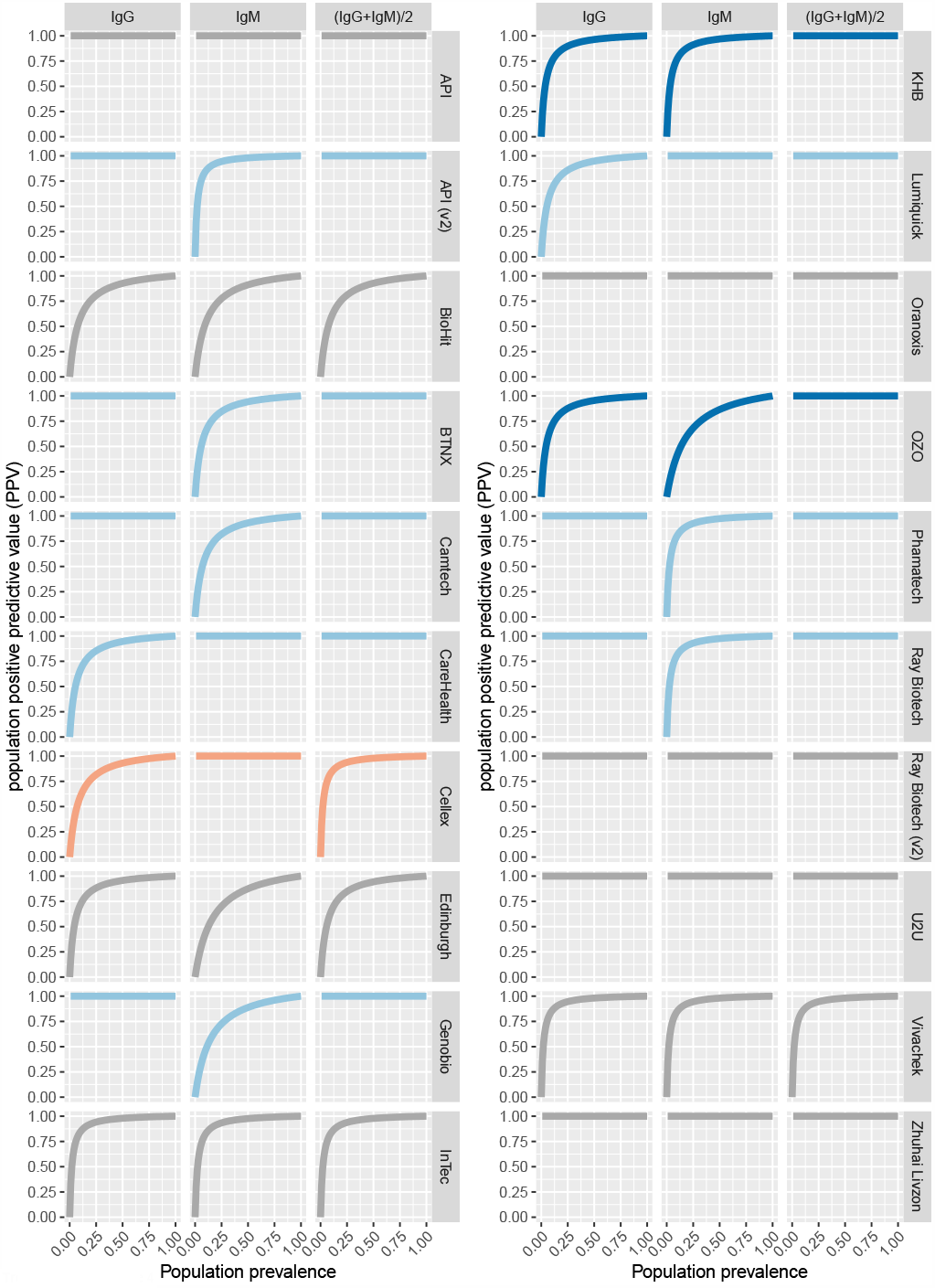
**Effect of a changing disease prevalence (x axis) on the population-level PPV of each test** for scores derived from IgG (left), IgM (middle), and the average score of both antibodies (right). As disease prevalence increases, the high burden on specificity of LFAs is reduced. Color coding: Grey: no improvement; Light blue: rescues one of the two antibodies; Dark blue: rescues both of the antibodies; Orange: performs worse than one of the antibodies.

To visualize the effect of changing population prevalence on the PPV, we created an interactive webapp (https://covid.omics.kitchen; **Figure 5)**, which allows the user to extrapolate the likelihood that they do in fact have SARS-CoV-2 antibodies if they have a positive LFA result, given the infection prevalence and test accuracy. The app includes benchmark performance of all 20 LFAs evaluated here, as well as those reported in Whitman et al^16^ (filtered to remove samples taken under 10 days post symptom-onset). In an effort to further generalize the utility of this tool, we allow the user to explore the effects of assay performance under difference prevalence scenarios, and to input the reported prevalence data from specific geographic locations within the US^1^, based on national, state, and county records.

**Figure 5:**
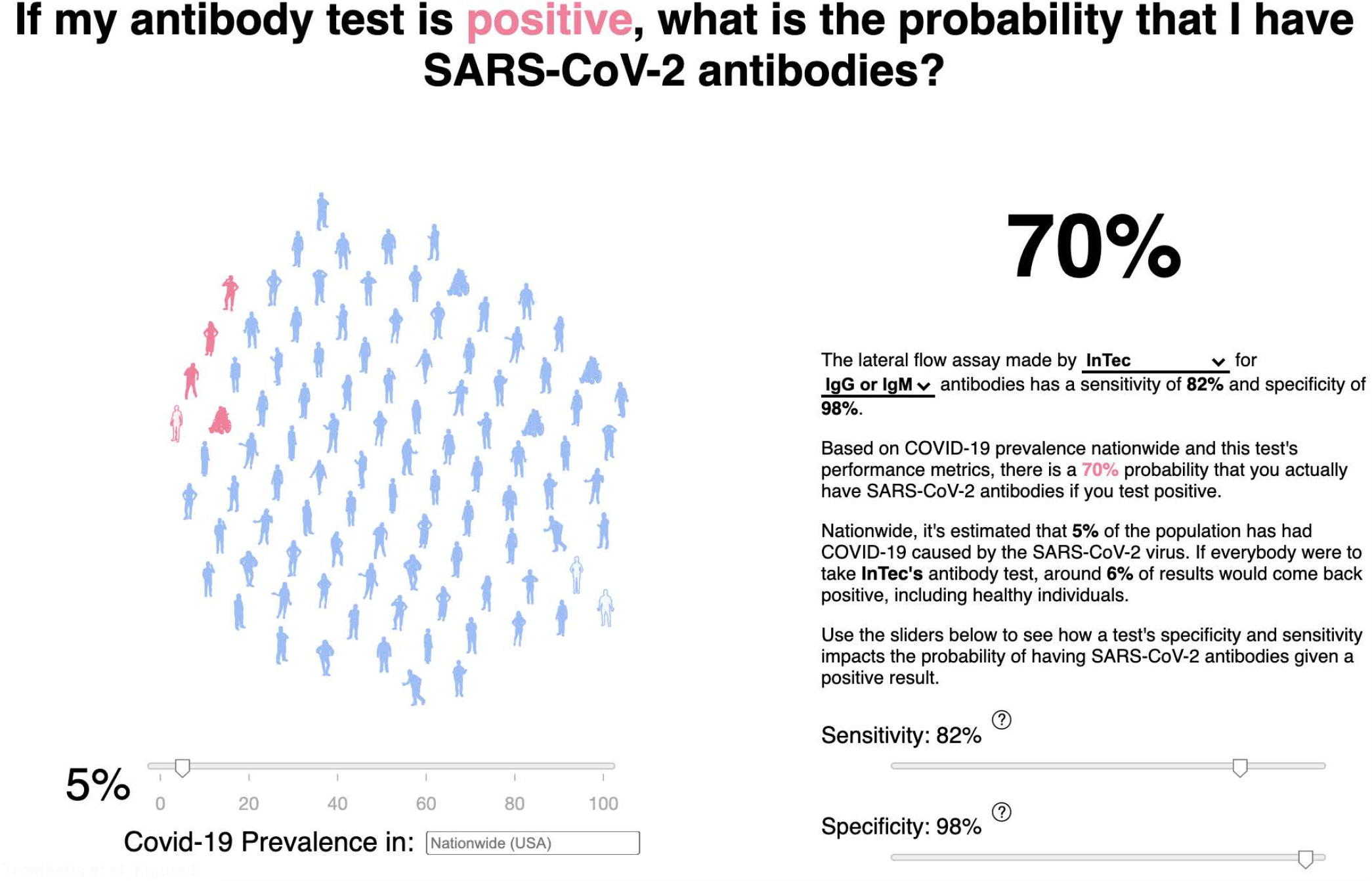
**Example image of the interactive webapp** hosted at https://covid.omics.kitchen. Using the sliders, one is able to visualize the effect of changing the disease prevalence (and the test performance) on the resulting probability that, given a positive LFA test, the individual does in fact have SARS-CoV-2 antibodies. The figure shows an example of positive predictive value using the US national prevalence of 5% (on November 27^th^, 2020) and the performance characteristics of the InTec LFA.

## Discussion

In this study, we report a standardized cross-evaluation of LFAs on the same pre-pandemic SARS-CoV-2-negative and PCR-confirmed SARS-CoV-2-positive samples, and rate their reproducibility, usability, and performance characteristics. Overall, the LFAs showed a higher propensity for false negative than false positive readings. Results are public: https://covidinnovation.partners.org/evaluation/. We use the Simoa technology^39^ to measure the concentrations of anti-spike protein IgG and IgM antibodies and extrapolate the assays’ limits of detection. We also established a web tool to aid users in understanding the likelihood they have anti-SARS-CoV-2 antibodies given a positive test result. This resource of performance characteristics of several LFAs and a tool for result interpretation, can both be used for immunosurveillance and future home testing applications.

LFAs are tractable tools to estimate community seroprevalence, especially with anticipated seasonal fluctuations in the transmission dynamics of SARS-CoV-2 and other viruses that cause the common cold, which confound the symptomatologic diagnosis of COVID-19^42^. As new waves of the COVID-19 pandemic resurge around the globe^1^, and with commencing vaccinations against SARS-CoV-2 infections^18, 19, 20, 22^, there is a renewed interest in serological tests to detect anti-SARS-CoV-2 antibodies^28, 43^. Affordable LFAs offer an attractive option for monitoring the presence and longevity of anti-SARS-CoV-2 antibodies, and determining population-level herd immunity^44^. LFAs also obviate the need for complex laboratories to process the samples^45^. As the pandemic expands to previously unexposed communities, it is critical to use simple tools to monitor exposure dynamics and seroconversion in SARS-CoV-2-exposed individuals, as well as vaccine-induced immunity^22^. We tested a mixture of LFAs targeting SARS-CoV2 nucleocapsid, spike proteins, or both. Moderna’s mRNA-1273 and Pfizer-BioNTech COVID-19 vaccines encode SARS-CoV2 spike proteins to induce anti-spike antibodies^20, 46^. Therefore, LFAs targeting the spike and nucleocapsid proteins of SARS-CoV2 could be used to differentiate vaccine- and infection-induced antibody responses, respectively.

Rigorous evaluation of these LFAs by manufacturer-independent parties is important. The US FDA independently reviews medical products before commercialization. The FDA used Emergency Use Authorization (EUA) authority to accelerate the implementation of diagnostic products during the pandemic. Commercial manufacturers were required to submit a completed EUA request^30^. Unfortunately, the rush to market introduced many tests that did not meet typical US or international standards^47^. Therefore, the FDA and international regulatory agencies continue to update guidelines for authorization of new serological tests. Our evaluation plan mirrored the FDA guidelines for evaluating serological tests^30^. We included 10 HIV-positive samples to test whether they have higher false positive results in SARS-CoV2-negative samples^48^, and did not detect higher false positive results.

The mere detection of IgG or IgM responses does not guarantee that neutralizing antibodies are present at protective titers ^13, 49^. The study demographics suggest a slight over-representation of African Americans among cases, as reported^50^. However, the sample size was underpowered to formally determine the effect of race on test performance. In our analysis, IgM detectability was less sensitive and reproducible than IgGs across multiple LFAs, possibly due to both lower IgM titres, and lower limits of detection for the assays. Waning antibody responses have been reported in some SARS-CoV-2-infected individuals^51, 52, 53, 54^. Furthermore, reported cases of re-infection with SARS-CoV-2 suggest that prior exposure, and even seroconversion, do not universally protect against SARS-CoV-2 infection^54, 55, 56^. This could result from low antibody titers as shown in an immunocompromised patient^57^, low durability of infection-induced antibodies^52, 53, 54^, or low neutralizing potential of SARS-CoV-2 antibodies in some individuals^44^. IgG and IgM antibodies may also target irrelevant epitopes outside the spike and receptor binding domains, and consequently be less efficient at intercepting infection by the virus^58^. The WHO cautions against interpreting presence of antibodies, even neutralizing ones, as lower risk of re-infection and transmission^59^. The presence of antibodies could, however, be used for rapid immunosurveillance to monitor extent of population transmission, particularly in asymptomatic but SARS-CoV2-exposed individuals^6, 43^.

One major concern about the deployment of these tests is the misinterpretation of positive results^13, 49^. As more tests move towards FDA clearance for home use, clear scientific communication about the result interpretation becomes more crucial^31,^ 49. A positive serological result does not necessarily mean active infection^31, 41, 60^. Although combined use of molecular and seroconversion results can be used to confirm the diagnosis of symptomatic and hospitalized individuals^45^, a positive serological test in the absence of symptoms dissociates the presence of the antibodies from the time of infection^54^. Additionally, it is important to understand the implication of false positive and false negative results, particularly in the context of a low-prevalence disease such as COVID-19^35^. Low prevalence decreases the negative predictive value of a test, but increases the rate of false positives^35^. A false positive serological test result may prematurely instill confidence that one has immunity against SARS-CoV-2 infection, thus resulting in behavioral changes that increase risk of transmission^61^. Hence, the probability that a person without antibodies will test negative on a serological test is more important than test sensitivity^59, 60, 61^.

Our study presents a few limitations. Although we successfully benchmarked the performance of the LFAs to a quantitative assay^39^, we did not determine the neutralizing potential of these antibodies. Secondly, samples were acquired when PCR testing was restricted to severely ill patients. For epidemiological studies and population surveillance, it will be important to evaluate assay performance on asymptomatic individuals.

In conclusion, our study provides a public resource to aid researchers, healthcare providers, public health professionals, and industries impacted by the pandemic such as airlines, in choosing the appropriate LFAs for their intended use cases.

## Data Availability

All evaluation data are available in a public repository: https://covidinnovation.partners.org/evaluation/
Web app to generate positive predictive values is available online: https://covid.omics.kitchen.

https://covid.omics.kitchen

## Acknowledgments

This study was possible through the Diagnostics Pillar of the Mass General Brigham COVID Center for Innovation (MGBCCI) (https://covidinnovation.partners.org/), predominantly staffed by volunteers. The LFA horizon scan and selection was performed by the Direct-to-Consumer working group, and the Diagnostics Accelerator and Validation working group developed the protocols and performed the evaluations. The study is supported by funding from the Massachusetts Consortium on Pathogen Readiness (MassCPR) (https://masscpr.hms.harvard.edu/).

## Online Supplement

### Supplementary Methods

#### Analysis

Computational analyses were performed in R/markdown. LFA evaluation reports are available (http://publicdata.omics.kitchen/Projects/MGBCCI/LFA/VendorReports/).

Formulae for computing performance metrics and confidence intervals are provided below. IgG/IgM bands were called via visual inspection by two experienced human operators. Invalid LFAs (e.g. missing a control band) were excluded entirely from further analysis and the sample was not re-run.

A score was computed for each sample/assay/antibody combination, using the following algorithm:

- Each of N operators reading the LFA were assigned a weight of 1/N to final score
- If an operator observed a band, the score would increase by 1/N. If the operator saw no band, the score would decrease by 1/N

This process was performed separately for the IgG and IgM channels, with an overall score produced for each antibody. This per-sample/per-antibody score thus has the following values: −1 (all operators agree: no band), 0 (operators disagree), and +1 (all operators agree: band observed). Given that COVID-19 remains, even in late 2020, a low-prevalence disease, we apply a conservative case definition (score ≥0), where discordant operator readings (score=0) are classed as negative for presence of the antibody, in order to favor specificity. A combined IgG and IgM score was computed as the average of the two individual scores: (IgG+IgM)/2.

Reproducibility analysis used the same scoring system above. Tests of the COVID+ pool that received a score >0 were called “correct”. COVID-tests were ones that received a score <0. A score of 1 or −1 was called “consistent”, with all other scores indicating disagreement between operators called as “inconsistent.” These two classifications were concatenated to produce the outcomes “correct consistent”, where both operators agreed on the correct outcome, “incorrect consistent,” where both operators agreed on the incorrect outcome, and “inconsistent,” where operators disagreed on the outcome. The outcome of each individual test was plotted as a proportion of total tests of each pool on each of the two days of testing.

#### Webapp

We developed an interactive web-application (https://covid.omics.kitchen) to help visualize false positive and false negative LFA readings based on test accuracy and disease prevalence. We incorporate data from the 20 LFAs reported here as well as the 6 LFAs evaluated by Whitman, et al^16^. County, State, and US-wide disease rates (cumulative numbers of individuals confirmed to have been infected since the start of the outbreak) are pulled dynamically from the New York Times github repository (https://github.com/nytimes/covid-19-data). The webapp is implemented in d3.js JavaScript and hosted on Amazon Web Services AWS/Amplify.

#### Formula

##### Sensitivity

- *n* = *TP* + *FN*
- 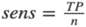
- 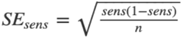
- 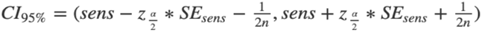

##### Specificity

- *n = TN* + *FP*
- 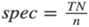
- 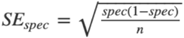
- 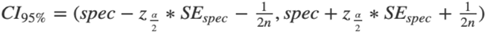

##### PPV

- *n = TP* + *FP*
- 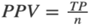
- 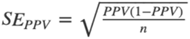
- 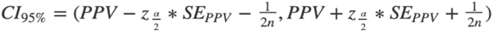

##### NPV

- *n = TN* + *FN*
- 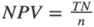
- 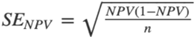
- 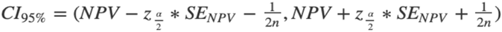

**Supplementary Table 1:** LFA commercial kit information, sample requirements, and protocol details

| Manufacturer | Lot Number | Biological Target | Sample Requirement | IgG/IgM Cassette Structure | Total input volume (uL) | Incubation time (minutes ) |
| --- | --- | --- | --- | --- | --- | --- |
| API_V1 | CoV1252004C | N and S | whole blood, plasma, serum | Same strip | 10 | 15-20 |
| API_V2 | COV1252008B | N and S | whole blood, plasma, serum | Same strip | 10 | 15-20 |
| BioHit | SA200401 | N and S | venous whole blood, plasma, serum | Same strip | 10 | 15-20 |
| BTNX | I2004027 | N and S | whole blood, plasma, serum | Same strip | 5 | 15 |
| Camtech | CAM240420 | S | plasma, serum | same strip | 10 | 15 |
| CareHealth | G070720 | N | venous whole blood, plasma, serum | Same strip | 5 | 10-15 |
| Cellex | 20200503WI5515C025 | N and S | venous whole blood, plasma, serum | Same strip | 10 | 15-20 |
| Edinburgh | 2000798A | N and S | whole blood (capillary and venous) | Same strip | 20 | 10 |
| Genobio | VMG200331 | N and S | venous whole blood, plasma, serum | Same cassette, separate strips | 20 | 10 |
| InTec | ITP6002-TC25/GJ20030288 | N and S | whole blood (venous and fingerstick), plasma, serum | Same cassette, separate strips | 20 | 15-20 |
| KHB | 423200332 | N | fresh whole blood, plasma, serum | Same strip | 10 | 15 |
| Livzon | CK2004240410 | N and S | venous whole blood, plasma, serum | Separate cassettes | 20 | 10-15 |
| Lumiquick | 2004219 | N and S | venous whole blood, plasma, serum | Same strip | 2 | 15 |
| Oranoxis | RC-0220 | S | plasma, serum | Same strip | 10 | 15 |
| Ozo | P2002 | S | whole blood (capillary and venous), plasma, serum | Same cassette, separate strips | 20 | 10 |
| Pharmatech | D00647 | N | whole blood (venous and fingerstick), plasma, serum | Same strip | 10 | 10 |
| RayBiotech_V1 | 501202955 | N | whole blood (venous and fingerstick), serum | Same cassette, separate strips | 25 | 10 |
| RayBiotech_V2 | 715202954 | N | whole blood (venous and fingerstick), serum | Same cassette, separate strips | 25 | 10 |
| U2U | 172004-01 | N and S | whole blood (venous and fingerstick), plasma, serum | Same strip | 10 | 15 |
| VivaCheck | SU2005001 | N and S | whole blood (venous and fingerstick), serum, heparin plasma | Same strip | 10 | 15 |

**Supplementary Table 2:** IFU clarity rubric. Kits were assigned one point for each criteria listed if met

|  | Criteria for IFU clarity |
| --- | --- |
| 1 | Includes explanation of intended use |
| 2 | Storage conditions of provided kit components are listed |
| 3 | Sample requirements and collection methods (e.g. whole blood, serum, plasma, equilibration temperature, etc.) |
| 4 | Storage information for specimens |
| 5 | Precautions/provides warnings and conditions to avoid (e.g. do not use expired cassettes, etc.) |
| 6 | Test procedure: temperature specified for running sample and cassette |
| 7 | Test procedure: clearly states when cassette should be read |
| 8 | Test procedure: clearly states valid time window (e.g. test results invalid after a certain interval of time) |
| 9 | Test procedure: provides visual diagrams for protocol steps |
| 10 | Provides instruction on how to proceed if results are invalid (e.g. repeat test, or increase incubation time) |
| 11 | Includes a rubric for interpretation of results |
| 12 | Provides visual diagrams or cassette images to assist with interpretation of results (has to include at least an example of a positive and negative readout) |
| 13 | Clearly describes correct volume to pipette (e.g. marked pipette, markings explained in the IFU, or dropwise measurement) |
| 14 | Complete instructions provided for lancets or other accessories included for use (e.g. instructions present for all kit components) |

**Supplementary Table 3.**
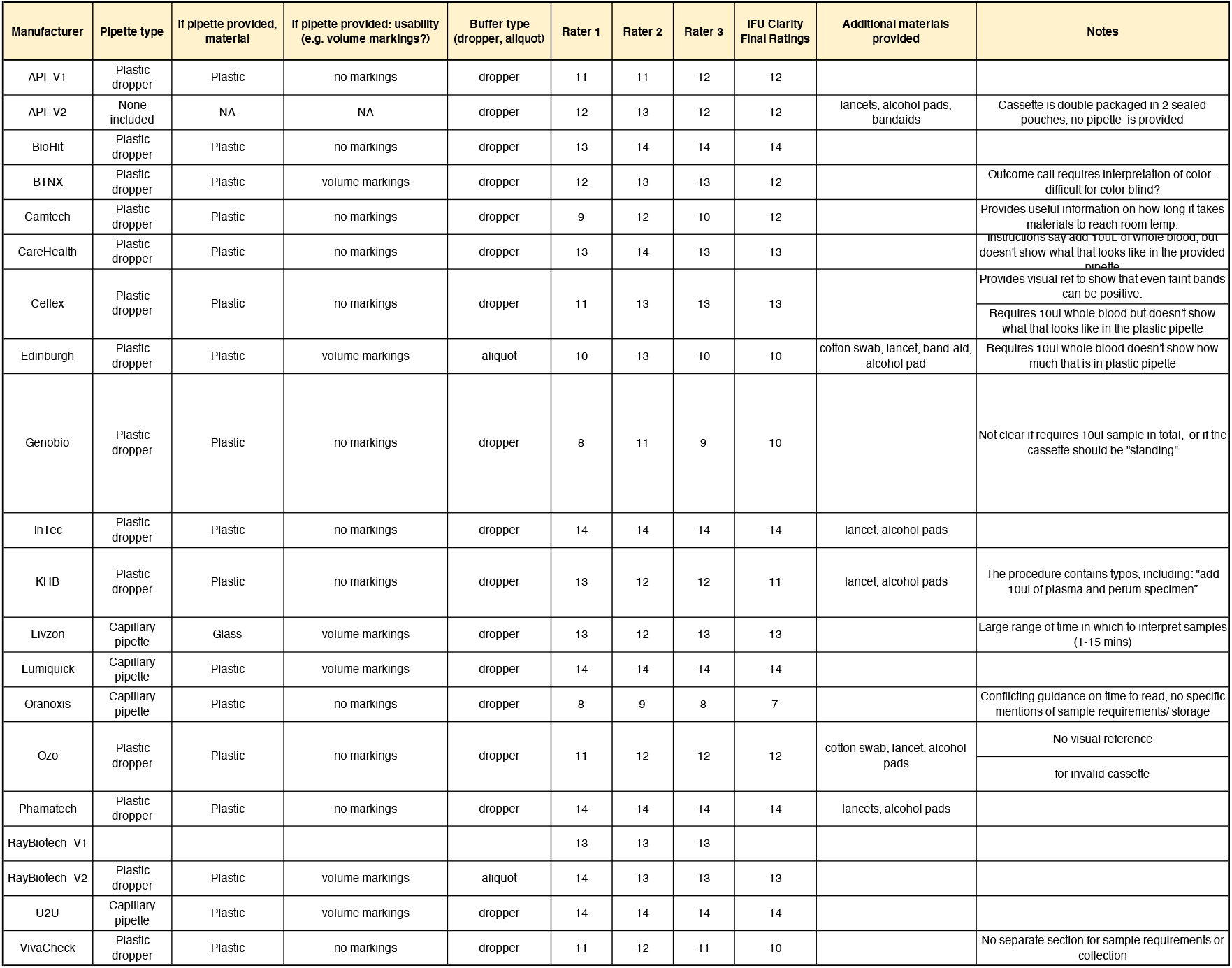
Usability ratings and description of kit components.

**Supplementary Table 4.**
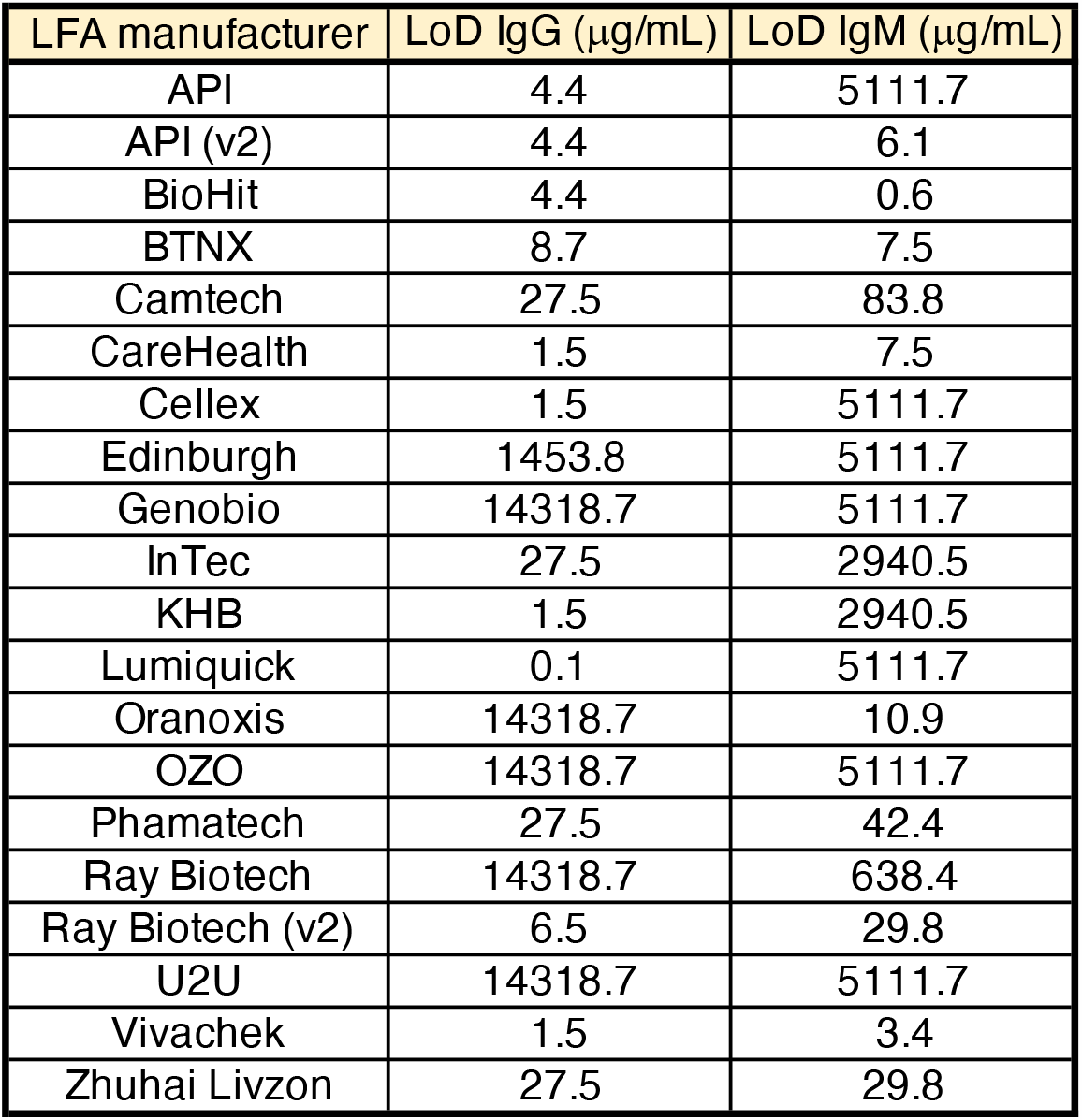
LFA Limits of detection estimated from anti-spike antibodies concentrations measured by Simoa; amounts reflect lowest antibody concentrations in plasma samples that show a positive band

## References

1. Dong, E., Du, H. & Gardner, L. An interactive web-based dashboard to track COVID-19 in real time. Lancet Infect Dis 20, 533–534 (2020).

2. Editors. Dying in a Leadership Vacuum. N Engl J Med 383, 1479–1480 (2020).

3. Looi, M.K. Covid-19: Is a second wave hitting Europe? BMJ 371, m4113 (2020).

4. Wiersinga, W.J., Rhodes, A., Cheng, A.C., Peacock, S.J. & Prescott, H.C. Pathophysiology, Transmission, Diagnosis, and Treatment of Coronavirus Disease 2019 (COVID-19): A Review. JAMA 324, 782–793 (2020).

5. Oran, D.P. & Topol, E.J. Prevalence of Asymptomatic SARS-CoV-2 Infection : A Narrative Review. Ann Intern Med 173, 362–367 (2020).

6. Havers, F.P. et al.. Seroprevalence of Antibodies to SARS-CoV-2 in 10 Sites in the United States, March 23-May 12, 2020. JAMA Intern Med (2020).

7. Ehrlich, H., Boneva, D. & Elkbuli, A. The intersection of viral illnesses: A seasonal influenza epidemic amidst the COVID-19 pandemic. Ann Med Surg (Lond) 60, 41–43 (2020).

8. Tahmasebi, S., Khosh, E. & Esmaeilzadeh, A. The outlook for diagnostic purposes of the 2019-novel coronavirus disease. J Cell Physiol 235, 9211–9229 (2020).

9. Kumar, R., Nagpal, S., Kaushik, S. & Mendiratta, S. COVID-19 diagnostic approaches: different roads to the same destination. Virusdisease 31, 97–105 (2020).

10. Hellou, M.M. et al.. Nucleic-acid-amplification tests from respiratory samples for the diagnosis of coronavirus infections: systematic review and meta-analysis. Clin Microbiol Infect (2020).

11. D’Cruz, R.J., Currier, A.W. & Sampson, V.B. Laboratory Testing Methods for Novel Severe Acute Respiratory Syndrome-Coronavirus-2 (SARS-CoV-2). Front Cell Dev Biol 8, 468 (2020).

12. Pecora, N.D. & Zand, M.S. Measuring the Serologic Response to Severe Acute Respiratory Syndrome Coronavirus 2: Methods and Meaning. Clin Lab Med 40, 603–614 (2020).

13. Peeling, R.W. et al.. Serology testing in the COVID-19 pandemic response. Lancet Infect Dis 20, e245–e249 (2020).

14. La Marca, A. et al. Testing for SARS-CoV-2 (COVID-19): a systematic review and clinical guide to molecular and serological in-vitro diagnostic assays. Reprod Biomed Online 41, 483–499 (2020).

15. Pickering, S. et al.. Comparative assessment of multiple COVID-19 serological technologies supports continued evaluation of point-of-care lateral flow assays in hospital and community healthcare settings. PLoS Pathog 16, e1008817 (2020).

16. Whitman, J.D. et al.. Evaluation of SARS-CoV-2 serology assays reveals a range of test performance. Nat Biotechnol 38, 1174–1183 (2020).

17. Oliver, S.E. et al.. The Advisory Committee on Immunization Practices’ Interim Recommendation for Use of Pfizer-BioNTech COVID-19 Vaccine - United States, December 2020. MMWR Morb Mortal Wkly Rep 69, 1922–1924 (2020).

18. Jackson, L.A. et al.. An mRNA Vaccine against SARS-CoV-2 - Preliminary Report. N Engl J Med 383, 1920–1931 (2020).

19. Anderson, E.J. et al.. Safety and Immunogenicity of SARS-CoV-2 mRNA-1273 Vaccine in Older Adults. N Engl J Med (2020).

20. Walsh, E.E. et al.. Safety and Immunogenicity of Two RNA-Based Covid-19 Vaccine Candidates. N Engl J Med (2020).

21. Dagotto, G., Yu, J. & Barouch, D.H. Approaches and Challenges in SARS-CoV-2 Vaccine Development. Cell Host Microbe 28, 364–370 (2020).

22. Poland, G.A., Ovsyannikova, I.G. & Kennedy, R.B. SARS-CoV-2 immunity: review and applications to phase 3 vaccine candidates. Lancet 396, 1595–1606 (2020).

23. Zohar, T. et al.. Compromised Humoral Functional Evolution Tracks with SARS-CoV-2 Mortality. Cell 183, 1508–1519 e1512 (2020).

24. Lucas, C. et al.. Kinetics of antibody responses dictate COVID-19 outcome. MedRxiv (2020).

25. Wang, H., Ai, J., Loeffelholz, M.J., Tang, Y.W. & Zhang, W. Meta-analysis of diagnostic performance of serology tests for COVID-19: impact of assay design and post-symptom-onset intervals. Emerg Microbes Infect 9, 2200–2211 (2020).

26. Caini, S. et al.. Meta-analysis of diagnostic performance of serological tests for SARS-CoV-2 antibodies up to 25 April 2020 and public health implications. Euro Surveill 25 (2020).

27. Lisboa Bastos, M. et al. Diagnostic accuracy of serological tests for covid-19: systematic review and meta-analysis. BMJ 370, m2516 (2020).

28. Moura, D.T.H. et al.. Diagnostic Characteristics of Serological-Based COVID-19 Testing: A Systematic Review and Meta-Analysis. Clinics (Sao Paulo) 75, e2212 (2020).

29. Gutierrez-Cobos, A. et al.. Evaluation of diagnostic accuracy of 10 serological assays for detection of SARS-CoV-2 antibodies. Eur J Clin Microbiol Infect Dis (2020).

30. FDA. EUA Authorized Serology Test Performance. United States Food and Drug Administration https://www.fda.gov/medical-devices/coronavirus-disease-2019-covid-19-emergency-use-authorizations-medical-devices/eua-authorized-serology-test-performance (2020).

31. Bermingham, W.H., Wilding, T., Beck, S. & Huissoon, A. SARS-CoV-2 serology: Test, test, test, but interpret with caution! Clin Med (Lond) 20, 365–368 (2020).

32. Long, Q.X. et al.. Antibody responses to SARS-CoV-2 in patients with COVID-19. Nat Med 26, 845–848 (2020).

33. Lou, B. et al.. Serology characteristics of SARS-CoV-2 infection after exposure and postsymptom onset. Eur Respir J 56 (2020).

34. Zhao, J. et al.. Antibody Responses to SARS-CoV-2 in Patients With Novel Coronavirus Disease 2019. Clinical infectious diseases : an official publication of the Infectious Diseases Society of America 71, 2027–2034 (2020).

35. Tenny, S. & Hoffman, M.R. Prevalence. StatPearls: Treasure Island (FL), 2020.

36. Theel, E.S. et al.. The Role of Antibody Testing for SARS-CoV-2: Is There One? J Clin Microbiol 58 (2020).

37. Jorfi, M. et al.. Diagnostic technology for COVID-19: comparative evaluation of antigen and serology-based SARS-CoV-2 immunoassays, and contact tracing solutions for potential use as at-home products. MedRxiv (2020).

38. Charlson, M.E., Pompei, P., Ales, K.L. & MacKenzie, C.R. A new method of classifying prognostic comorbidity in longitudinal studies: development and validation. J Chronic Dis 40, 373–383 (1987).

39. Norman, M. et al.. Ultrasensitive high-resolution profiling of early seroconversion in patients with COVID-19. Nat Biomed Eng (2020).

40. Tian, X. et al.. Potent binding of 2019 novel coronavirus spike protein by a SARS coronavirus-specific human monoclonal antibody. Emerg Microbes Infect 9, 382–385 (2020).

41. Hernaez, R. & Thrift, A.P. High Negative Predictive Value, Low Prevalence, and Spectrum Effect: Caution in the Interpretation. Clin Gastroenterol Hepatol 15, 1355–1358 (2017).

42. Kissler, S.M., Tedijanto, C., Goldstein, E., Grad, Y.H. & Lipsitch, M. Projecting the transmission dynamics of SARS-CoV-2 through the postpandemic period. Science 368, 860–868 (2020).

43. Alter, G. & Seder, R. The Power of Antibody-Based Surveillance. N Engl J Med 383, 1782–1784 (2020).

44. Neagu, M. The bumpy road to achieve herd immunity in COVID-19. J Immunoassay Immunochem, 1–18 (2020).

45. Nilles, E.J. et al.. Evaluation of two commercial and two non-commercial immunoassays for the detection of prior infection to SARS-CoV-2. medRxiv (2020).

46. Corbett, K.S. et al.. SARS-CoV-2 mRNA vaccine design enabled by prototype pathogen preparedness. Nature 586, 567–571 (2020).

47. Badnjevic, A., Pokvic, L.G., Dzemic, Z. & Becic, F. Risks of emergency use authorizations for medical products during outbreak situations: a COVID-19 case study. Biomed Eng Online 19, 75 (2020).

48. Tan, S.S., Chew, K.L., Saw, S., Jureen, R. & Sethi, S. Cross-reactivity of SARS-CoV-2 with HIV chemiluminescent assay leading to false-positive results. J Clin Pathol (2020).

49. Gill, D. & Ponsford, M.J. Testing for antibodies to SARS-CoV-2. BMJ 371, m4288 (2020).

50. Kullar, R. et al.. Racial Disparity of Coronavirus Disease 2019 in African American Communities. J Infect Dis 222, 890–893 (2020).

51. Long, Q.X. et al.. Clinical and immunological assessment of asymptomatic SARS-CoV-2 infections. Nat Med 26, 1200–1204 (2020).

52. Ibarrondo, F.J. et al.. Rapid Decay of Anti-SARS-CoV-2 Antibodies in Persons with Mild Covid-19. N Engl J Med 383, 1085–1087 (2020).

53. Kutsuna, S., Asai, Y. & Matsunaga, A. Loss of Anti-SARS-CoV-2 Antibodies in Mild Covid-19. N Engl J Med 383, 1695–1696 (2020).

54. Milani, G.P. et al.. Serological follow-up of SARS-CoV-2 asymptomatic subjects. Sci Rep 10, 20048 (2020).

55. To, K.K. et al.. COVID-19 re-infection by a phylogenetically distinct SARS-coronavirus-2 strain confirmed by whole genome sequencing. Clinical infectious diseases : an official publication of the Infectious Diseases Society of America (2020).

56. Cohen, J.I. & Burbelo, P.D. Reinfection with SARS-CoV-2: Implications for Vaccines. Clinical infectious diseases : an official publication of the Infectious Diseases Society of America (2020).

57. Mulder, M. et al.. Reinfection of SARS-CoV-2 in an immunocompromised patient: a case report. Clinical infectious diseases : an official publication of the Infectious Diseases Society of America (2020).

58. Hussain, A. et al.. Targeting SARS-CoV2 Spike Protein Receptor Binding Domain by Therapeutic Antibodies. Biomed Pharmacother 130, 110559 (2020).

59. WHO. “Immunity passports” in the context of COVID-19. World Health Organization commentaries https://www.who.int/news-room/commentaries/detail/immunity-passports-in-the-context-of-covid-19. (2020).

60. Waller, J., Rubin, G.J., Potts, H.W.W., Mottershaw, A.L. & Marteau, T.M. ‘Immunity Passports’ for SARS-CoV-2: an online experimental study of the impact of antibody test terminology on perceived risk and behaviour. BMJ Open 10, e040448 (2020).

61. Brown, R.C.H., Kelly, D., Wilkinson, D. & Savulescu, J. The scientific and ethical feasibility of immunity passports. Lancet Infect Dis (2020).

